# Understanding Bilateral Motor Coordination in Stroke Using the Towel Folding Task: An Exploratory Biomechanical Study

**DOI:** 10.1101/2024.09.03.24313027

**Authors:** Jingyi Wu, Patrick Wai-Hang Kwong, Ananda Sidarta, Jack Jiaqi Zhang, Jingwen Zhuang, Yining Li, Kenneth NK Fong

## Abstract

**Objectives:** Coordination deficits in bilateral upper limbs make daily activities more difficult for stroke survivors. Previous studies showed worse kinematics during unilateral tasks compared to healthy individuals, but this was unclear for bimanual tasks. We aim to assess the potential of the towel folding task from the Wolf Motor Function Assessment as a measure of bimanual control by examining kinematic differences between stroke survivors and healthy individuals and correlating these differences with clinical parameters in the stroke group.

**Methods:** This was a cross-sectional design. Seventeen people with stroke and sixteen healthy individuals participated. Vicon motion capture obtained kinematics of bilateral upper limbs during the task, including movement time, initiation delay, velocity, trunk displacement, smoothness, and inter-/intra-limb coordination. Statistical analyses compared groups and correlated kinematic variables with clinical parameters.

**Results:** Stroke survivors had longer movement times (*P* < .001, Cohen’s d = 1.396), slower initiation (*P* < .001, Cohen’s d = 0.797), lower max velocity (*P* = .026, Cohen’s d = −.815; *P* < .001, Cohen’s d = −2.156; and *P* = .005, Cohen’s d = −.736; respectively), greater trunk displacement (*P* < .001, Cohen’s d = 2.173 and *P* < .001, Cohen’s d = 1.727, respectively), less smoothness (*P* = .031, Cohen’s d = 0.883 and *P* < .001, Cohen’s d = .725, respectively), and altered inter-/intra-limb coordination. Regarding bilateral elbow-elbow coordination, stroke group exhibited decreased in-phase patterns (*P* < .001, partial η² = .368) and increased anti-phase and non-hemiplegic elbow dominancy patterns (*P* = .001, partial η² = .298 and *P* = .004, partial η² = .244, respectively). Regarding bilateral shoulder-shoulder coordination, stroke group showed decreased hemiplegic shoulder leading patterns (*P* = .010, partial η² = .196) and increased anti-phase and non-hemiplegic shoulder dominancy patterns (*P* = .001, partial η² = .315 and *P* < .001, partial η² = .463, respectively). For hemiplegic shoulder-elbow coordination, stroke group showed decreased anti-phase patterns (*P* < .001, partial η² = .382) and increased elbow dominancy *P*atterns (*P* < .001, partial η² = .324). Fugl-Meyer Assessment scores positively correlated with smoothness and hemiplegic shoulder-elbow coordination (r = −.500, *P* = .039 and r = .600, *P* = .010, respectively), while Action Research Arm Test scores negatively correlated with movement initiation delay (r = −.600, *P* = .010).

**Conclusions:** This study enhances understanding of the folding towel task and may provide metrics to quantify bilateral coordination task performance in stroke survivors.

## Introduction

The deficit in the upper extremities’ motor function is the most common sequelae resulting from stroke, which limits patients’ activities of daily living abilities, results in permanent disability, and therefore brings a heavy burden to family and society (1). Recently, the majority of studies focused on the hemiplegic limbs and aimed to promote independence in activities of daily living (ADL) by improving the motor function of the hemiplegic limb (2–4). However, most daily activities deeply rely on the cooperation and coordination of bilateral upper extremities, such as operating the bottle and folding the towel. Some studies used motion capture to demonstrate movement quality in the non-hemiplegic upper limb was impaired, with respect to the healthy control (5), which may be partly explained through the corticospinal motor tract (CST) that remains 10-15% of fibers descend the same side of the cortical area between bi-hemispheres through the corpus callosum (6). In addition, compensatory strategies including the overuse of the trunk and non-hemiplegic upper limb after stroke have developed to assist perform ADL. Most importantly, coordination deficits in bilateral arms, including but not limited to impairments in smoothness and inter-/intra-limb coordination, have been observed in stroke patients, which make them complete everyday programs more difficult (7–11). Therefore, it is necessary to explore the changes in upper-limb bimanual coordination patterns after stroke for the advancement of an effective rehabilitation program.

Dysfunction in upper-limb bimanual coordination can be assessed using clinical scales like the Wolf Motor Function Test (WMFT). The WMFT includes 17 items that evaluate strength and task performance based on performance time and functional ability (12, 13). Of the WMFT items, folding a towel assesses upper-limb bimanual coordination by having the client grasp, and then fold the towel in half. However, clinical assessments face significant floor and ceiling effects (14) and are not sensitive enough to identify compensatory strategies like trunk displacement and overuse of the non-hemiplegic limb (7). They also fail to capture motor quality variables, such as smoothness and inter/intra-limb coordination, due to the use of ordinal scales (15).

Kinematic assessments using motion capture systems overcome these disadvantages by providing objective parameters that capture movement quality and monitor compensatory movements without floor or ceiling effects (16). Recent studies have shown that features extracted from motion capture can clearly distinguish between healthy individuals and stroke patients during various functional movements (reaching, drinking water) and correlate with the severity of stroke-specific impairments (17–26). The Second Stroke Recovery and Rehabilitation Roundtable highlighted the benefits of using kinematic variables to assess upper extremity function in stroke patients during functional tasks (27). However, most studies focus on the hemiplegic upper limb, with no research exploring the feasibility of using kinematic data and motion capture to quantify bimanual coordination tasks in people with stroke. A standardized and widely used, folding towel could be valuable for assessing bilateral coordination. A comprehensive assessment of spatiotemporal variables and movement quality is essential for identifying specific issues with bimanual coordination in stroke patients (10, 11). This approach allows for targeted, evidence-based plans to improve coordinated performance.

Based on the above, this study aimed to (1) use kinematic data from optical motion capture to quantify the movement quality of the towel folding, (2) explore the differences in kinematic parameters between stroke patients and healthy individuals during this bimanual task, and (3) determine the relationship between kinematic variables from towel folding and upper limb impairment or activity limitations assessed by clinical scales in people with stroke. This approach aims to provide precise rehabilitation interventions and quantitative assessments of treatment outcomes.

## Materials and Methods

### Experimental design

This cross-sectional study enrolled a total of 33 participants, with 17 people with stroke and 16 healthy individuals between the ages of 18 and 75 years. Participants were recruited from local self-help organizations for stroke survivors from March 2022 to December 2022. Ethical approval was obtained from the Institute Review Board of the Hong Kong Polytechnic University (HSEARS20220125002). Written informed consent was obtained for all study participants enrolled in the study. The study was conducted in the university motion capture laboratory.

### Inclusion and exclusion criteria

All participants with stroke met all of the following criteria: (1) between 18 and 75 years of age, (2) diagnosed as first-ever stroke, confirmed by neuroimaging examination, (3) with mild or moderate motor impairment by Upper Limb Fugl Meyer Assessment (31 < UE-FMA < 66) (19), (4) with post-acute stroke (> 3 months after the ictus) (28); (5) able to follow instructions and carry out the experimental procedure with both arms. All healthy controls met all of the following criteria: (1) between 18 and 75 years of age, (2) right-hand dominant, (3) without a history of epilepsy or other psychiatric episodes.

All participants who met any of the following criteria were excluded: (1) current use of central neural system-affect medications, (2) other diseases substantially affecting the motor function of upper extremities and hands.

### Experimental setup and procedure

This study used a Vicon’s motion system with nine infrared cameras operating at 100 Hz to track the 3D movements of 50 reflective markers placed on a participant’s trunk and bilateral upper limbs (Fig 1). Visual3D software (C-Motion, Rockville, MD, USA) was utilized to create a scaled human model for each participant. The movement analysis was based on an X-Y-Z coordinate system, where X represented lateral-to-medial direction, Y represented anterior-to-posterior direction, and Z represented vertical direction.

**Fig 1.**
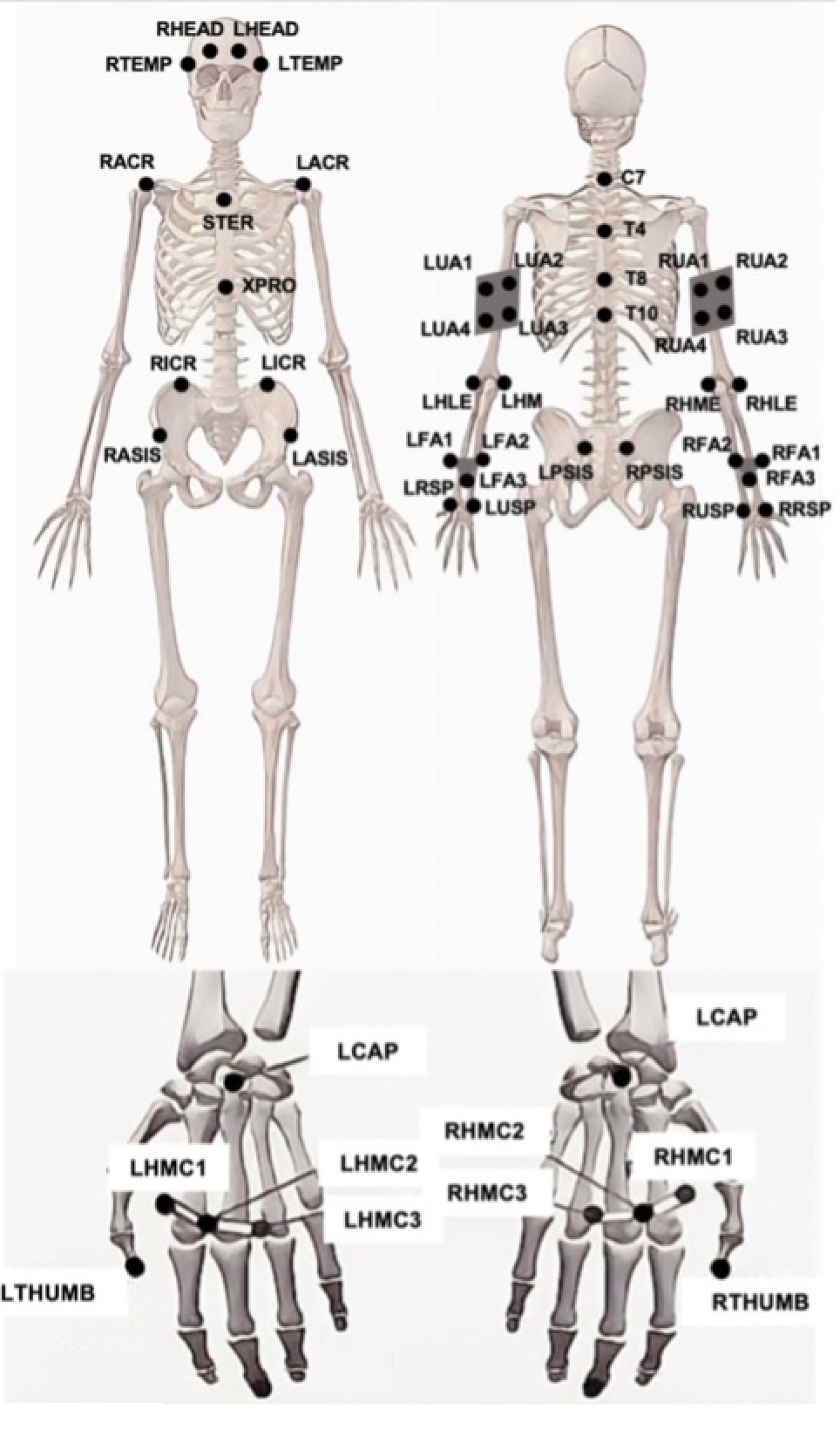
Markers placement

Participants were seated without back support and performed a towel-folding task following specific steps:

1. First, they placed both arms on the table with hands flat, shoulders neutral, and elbows bent (Fig 2a). Next, they reached to grasp the towel’s corners and folded it front to back (Fig 2b). Then, one hand (hemiplegic for participants with stroke, non-dominant for healthy individuals) grasped the towel’s corners while the other returned to the initial position (Fig 2c). Following this, the towel was folded from side to side using the hemiplegic hand (Fig 2s). Finally, the hemiplegic hand returned to the initial position (Fig 2e).
2. A total of 5 trials was recorded, with 20 seconds between each trial, ensuring a self-paced and standardized procedure for data collection.

**Fig 2.**
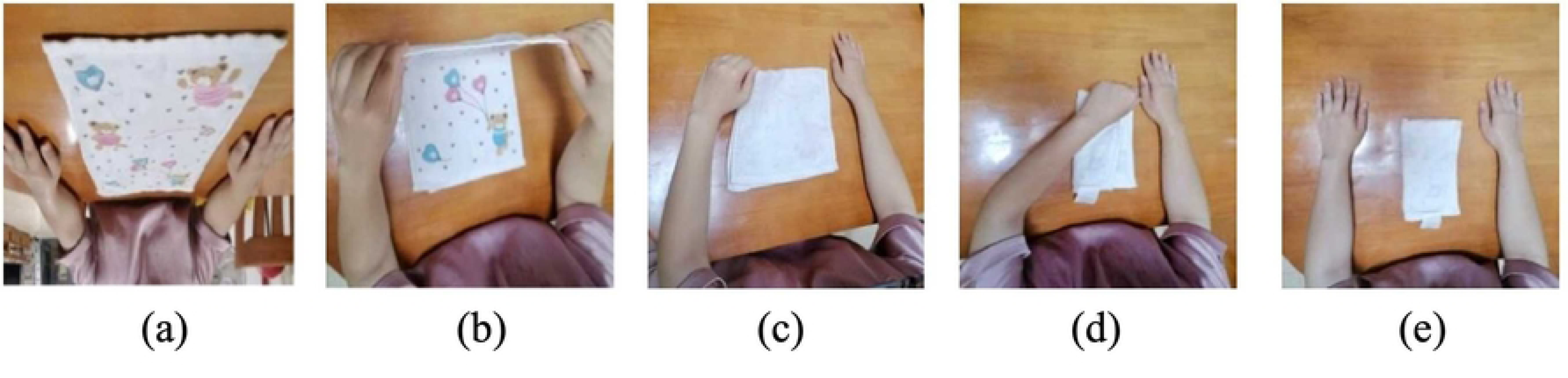
**Steps of folding the towel**

### Outcome measures

The Upper Extremity Fugl-Meyer Assessment (UE-FMA) and the Action Research Arm Test (ARAT) were used for standardized assessments in people with stroke.

The UE-FMA is a validated measure of movement impairment in stroke individuals (29, 30). The scale consists of 33 items to assess movements, reflexes, and coordination of the shoulder, elbow, forearm, wrist, and hand. The UE-FMA ranges from 0-66 points, with a maximum score of 66 points indicating the least motor deficit, with a score >31 indicating moderate or mild upper limb impairment (19).

The ARAT is a validated measure of upper limb functional limitations and activity levels in stroke individuals (31, 32). The scale comprises 19 items where 0 indicates no movement and 3 indicates normal movement. Items are categorized into 4 subscales (grasp, grip, pinch, and gross movement) to assess upper extremity performance of coordination, dexterity, and functioning. The ARAT total score ranges from 0 to 57, with a higher score indicating better performance (33).

### Data reduction and analysis

Reflective markers were labeled, and missing data were gap-filled using Nexus software (version 2.5). Visual 3D software (C-Motion, Rockville, MD, USA) was used to perform biomechanical modeling. Joint angles and joint velocity were extracted for further analysis. VectorCodeR package was used to calculate the coupling angle (CA) in the sagittal plane. Definitions of different kinematics variables were represented in detail in Table 1.

**Table 1.**
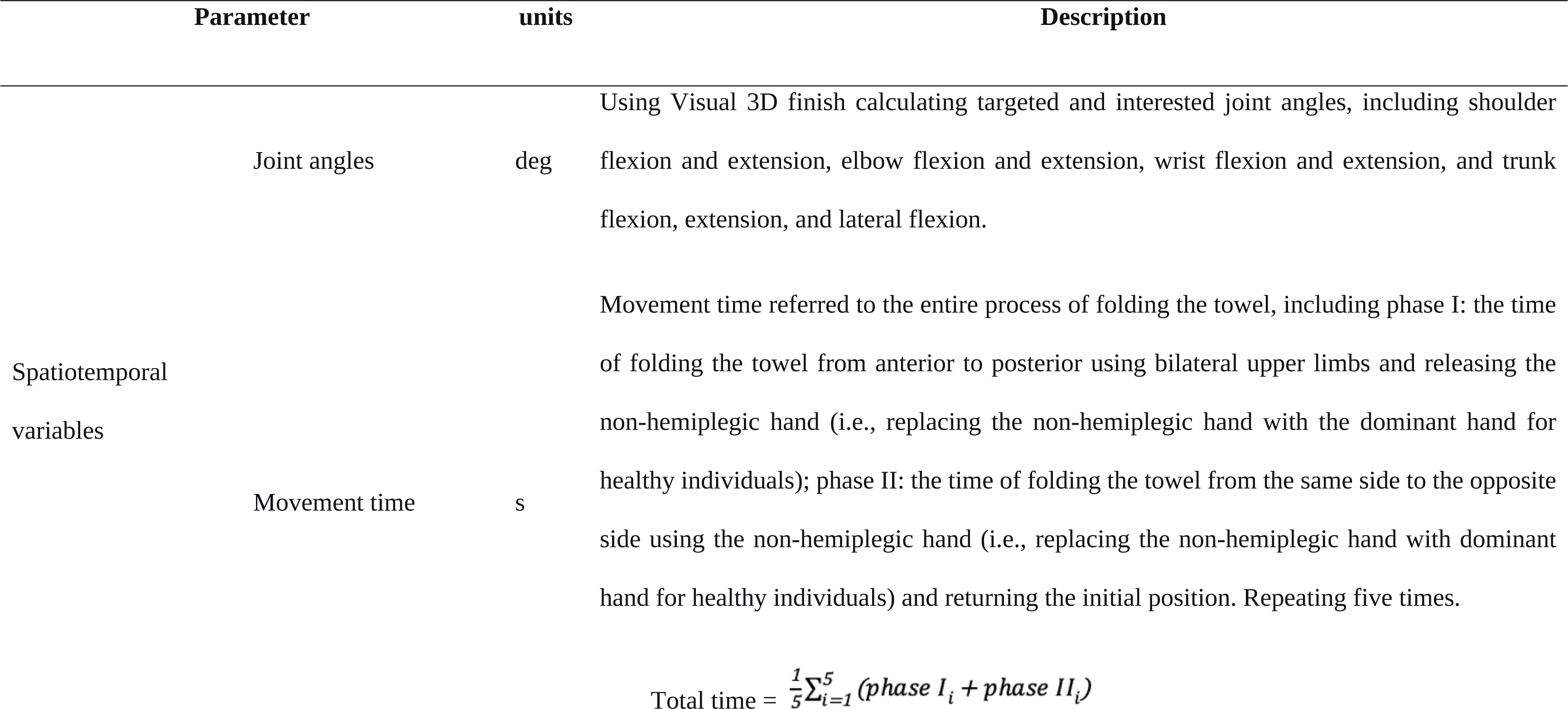
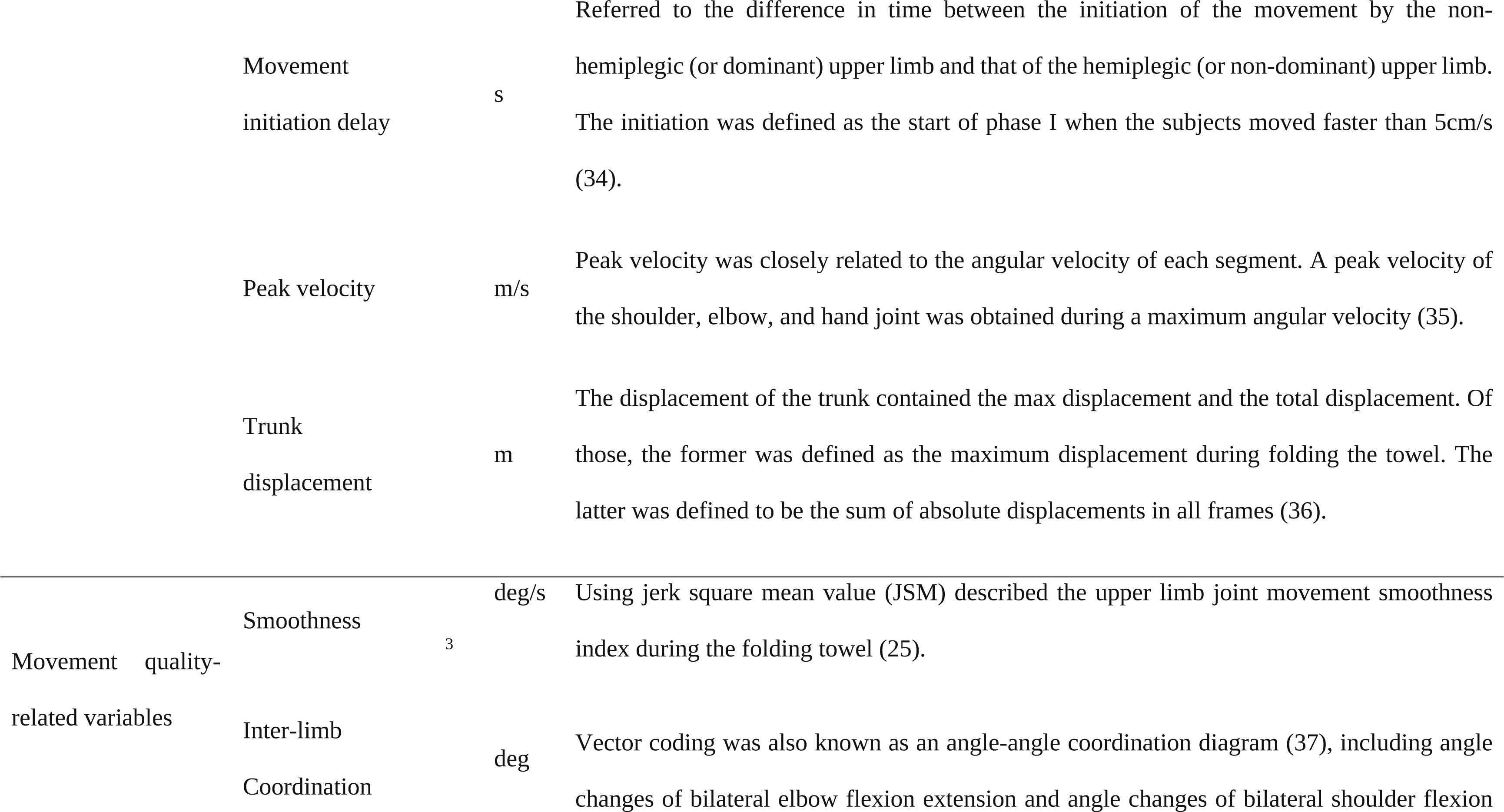
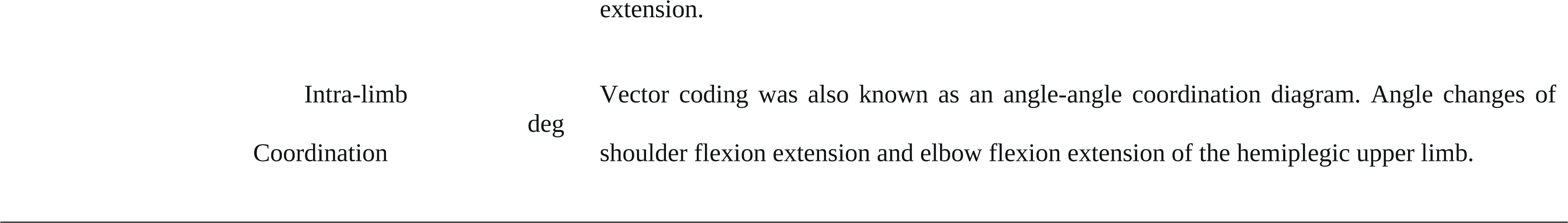
Definitions of kinematics variables used in the study.

**Peak velocities** were determined based on the whole process of the task as follows:

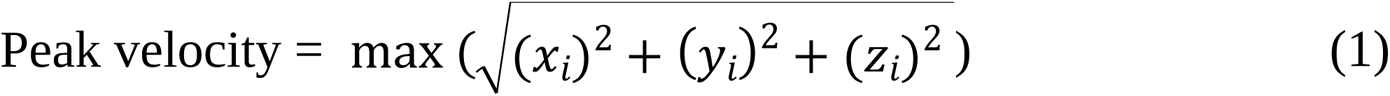

where *x*_*i*_, *y*_*i*_, and *z*_*i*_ are the coordinates of a marker, and i is the index of a frame.

**The total displacement of the trunk** was defined as the sum of absolute displacements in all frames as follows:

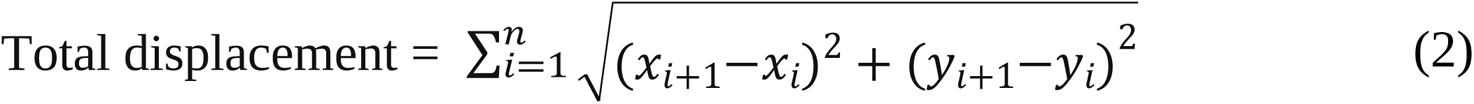

**Maximum displacement of the trunk** was defined as the maximum absolute displacement in all frames as follows:

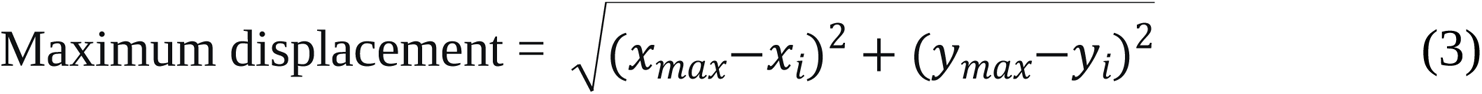

where *x*_*i*_ and *y*_*i*_ were the coordinates of a marker, and i was the index of a static frame.

**Smoothness** was quantified using the jerk square mean value (JSM) (25). JSM served as the average of the square of the joint angle third derivative value, according to the jerk third derivative of the position data as follows. Each subject’s JSM values were calculated for the shoulder, elbow, and wrist joints. Movement smoothness seemed to decrease when JSM increased:

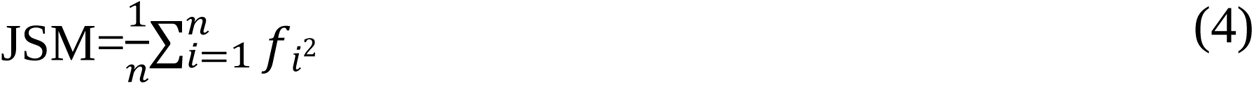

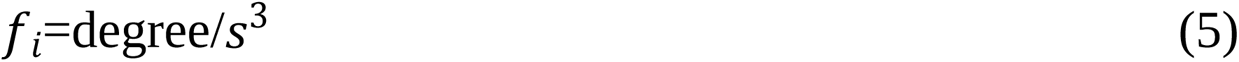

*f*_*i*_ represented the jerk value of joint angles that were calculated by the third derivative of the joint angles. n served as the number of data points.

**Coordination** between and within limbs was quantified using the vector coding techniques described by Needham et al. (37, 38). Vector coding measured the continuous interaction between two adjacent segments by analyzing the vector orientation between consecutive data points on the angle-angle diagram. This vector orientation, known as the coupling angle (CA), ranges from 0-360°. This study focused on bilateral elbow flexion-extension, bilateral shoulder flexion-extension, and hemiplegic shoulder and elbow flexion-extension couplings. Each coordination included four patterns (39).

For each instant (i) during folding the towel cycle, the coupling angles (γi) were obtained based on the consecutive proximal segment angles (shoulder) or hemiplegic side angle (non-dominant for healthy individuals) (θ_P(i)_, θ_P(i+1)_) and consecutive distal segment angles (elbow) or non-hemiplegic side (dominant for healthy individuals) (θ_D(i)_, θ_D(i+1)_) as follows:

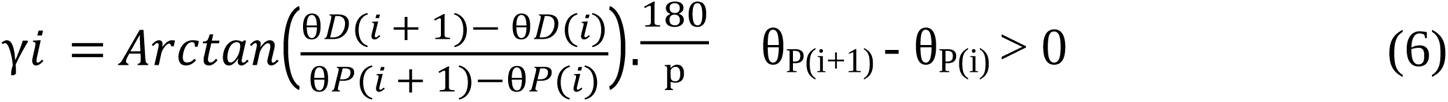

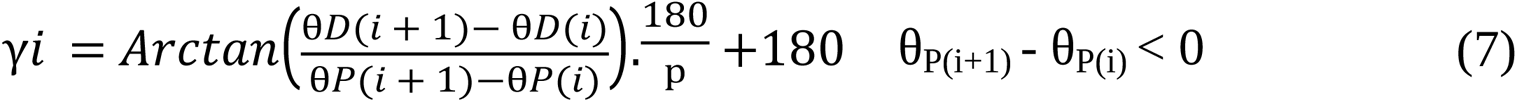

The following conditions were applied as follows:

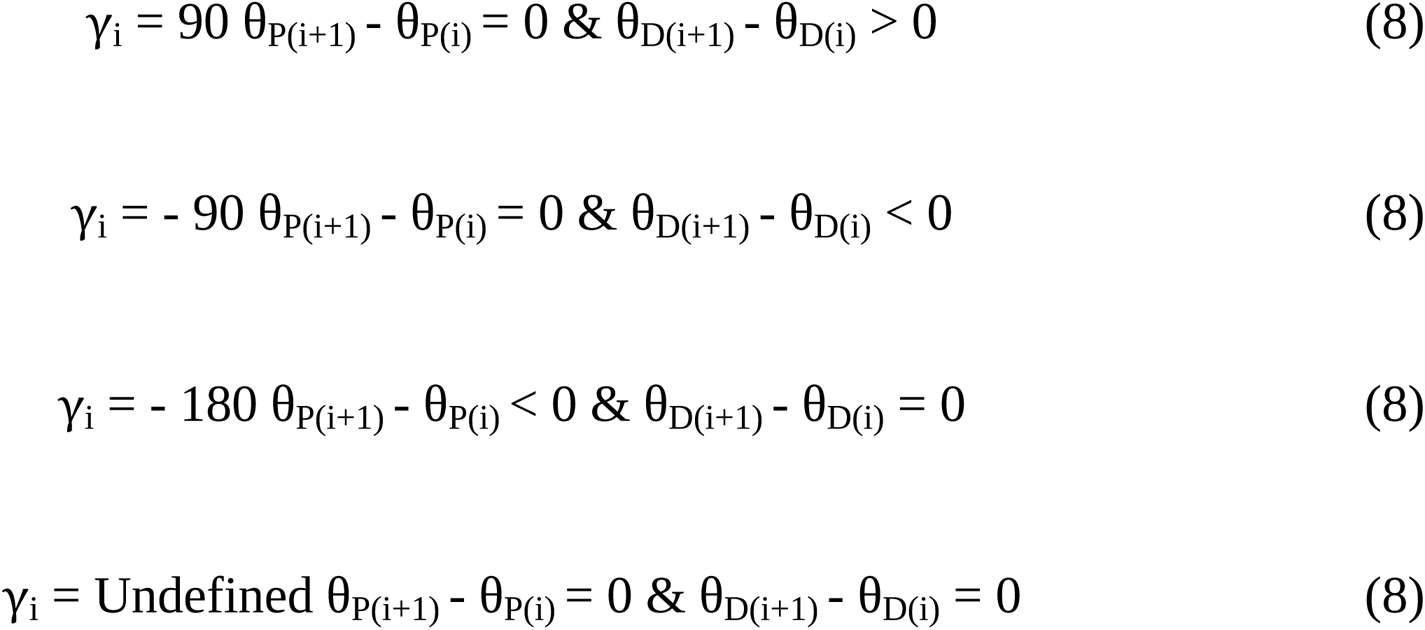

The coupling angle (γi) was corrected to present a value between 0 and 360 as follows (37, 40).

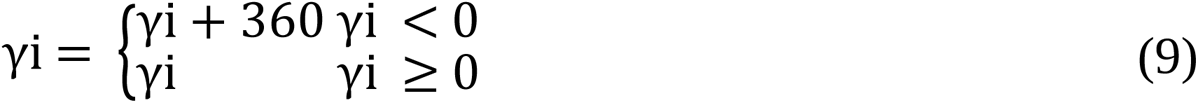

For ease of analysis and visualization, the similarity among three coordination patterns (bilateral elbow-elbow, bilateral shoulder-shoulder, and hemiplegic shoulder-elbow) between stroke and healthy individuals was calculated using the unit vector scalar product (DOT). DOT was maximum when the angles between two vectors were 0° (Fig 3). DOT was zero when the angle between two vectors was 90° as follows:

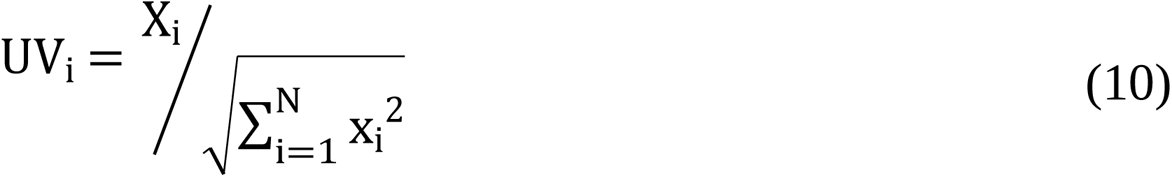

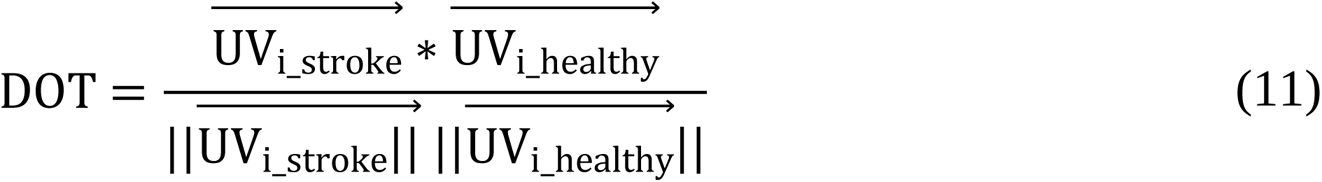

**Fig 3.**
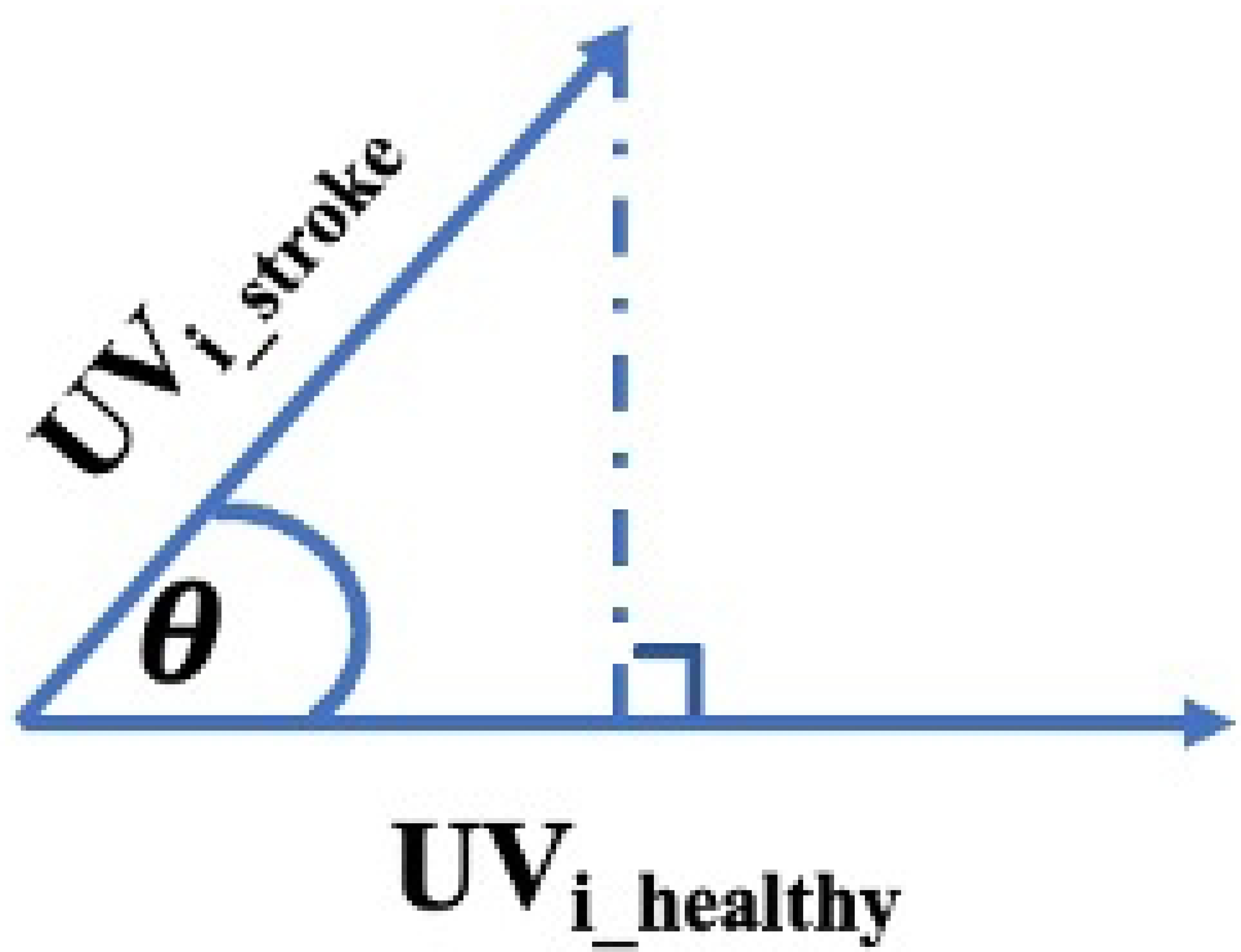
**DOT between individuals with stroke and health.**

X_i_ is the *i*th percentage of N basic vectors.

### Statistical analysis

R software v.4.2.2 (R Development Core Team, 2010) served as performing data analysis. The normality of the distribution of all variables was tested using Shapiro-Wilk’s test (41). Two-independent sample t-test, Mann–Whitney *U* test, and chi-squared test to examine the differences in demographic, kinematic variables, and clinical parameters between the two groups.

To determine whether the mean differences in inter-/intra-limb coordination (each coordination included four patterns) were significant between the two groups, a multivariate analysis of variance (MANOVA) was performed. Multiple comparisons were performed via Bonferroni adjustments at *p* < 0. 0125.

Pearson’s r correlation or Spearman rank order correlation was applied to test the correlation between kinematic variables and clinical parameters, depending on the data distribution. The statistics level was set at p<0.05. The effect size of the difference between groups was calculated by Cohen’s d and partial eta squared, where 0.2, 0.5, 0.8, and 0.01, 0.06, and 0.14 demonstrated small, medium, and large effect sizes, respectively, and the corresponding values for r were 0.1, 0.3, and 0.5 (42).

## Results

### Participants characteristics

Seventeen post-acute stroke individuals with mild-moderate motor impairment and sixteen right-handed healthy individuals were enrolled. There were no significant differences in age and gender between the two groups (Table 2).

**Table 2.** Demographic data and clinical assessments between stroke and healthy group.

| Demographic data | Stroke Group<br>(n = 17) | Healthy Group<br>(n = 16) | *P* value |
| --- | --- | --- | --- |
| Age | 63.0 (5.3) | 62.3 (6.3) | 0.892 |
| Sex (M/F) | 9/8 | 9/7 | 1.000 |
| Hemiplegic side/Handedness<br>(R/L) | 7/11 | R:16; L:0 | NA |
| Post-stroke duration (month) | 4.6 (1.9) | NA | NA |
| **Clinical parameters** |  |  |  |
| UE-FMA | 43.0 (19.0) | NA | NA |
| ARAT | 50.0 (28.0) | NA | NA |

Mean (SD); SD: standard deviation; M: male; F: female; R: right; L left.
UE-FMA: Upper Extremity Fugl-Meyer Assessment; ARAT: Action research arm test.
Median (IQR); IQR: interquartile range

### Exploring Kinematic Variables and Clinical Parameters

#### Time and Velocity

Compared to the healthy group, the stroke group had significantly longer movement time (mean difference = 27.9 s; *P <* .001; Cohen’s d = 1.396) and movement initiation delay (mean difference = .3 s; *P <* .001; Cohen’s d = .797) while folding the towel. In addition, compared with the healthy group, the stroke group had significantly lower peak velocities on non-hemiplegic elbow and bilateral shoulder joints (Tables 3 and 4). We found no notable differences in hemiplegic elbow velocity of stroke patients and non-dominant elbow velocity of healthy controls (Table 4).

**Table 3.**
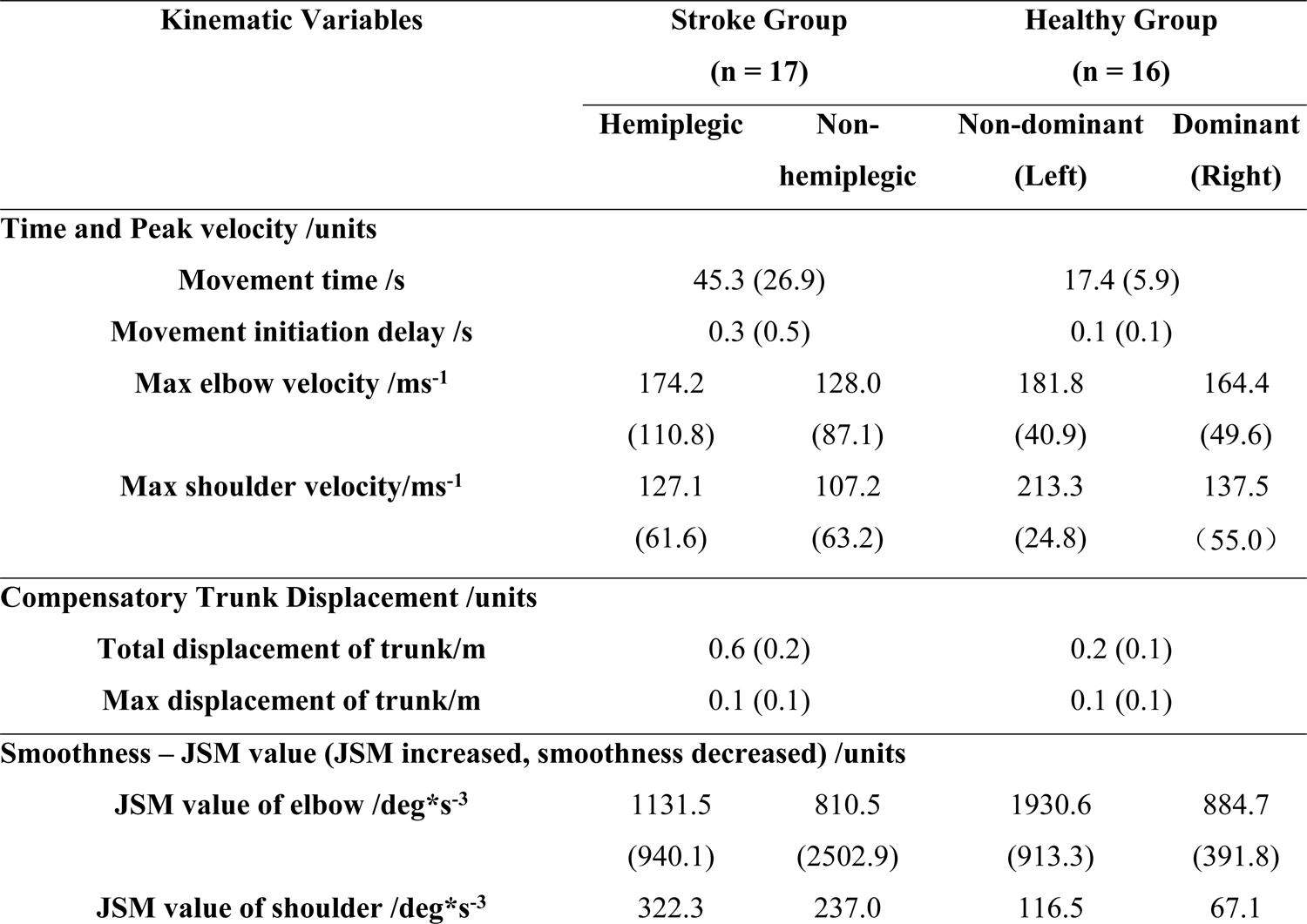
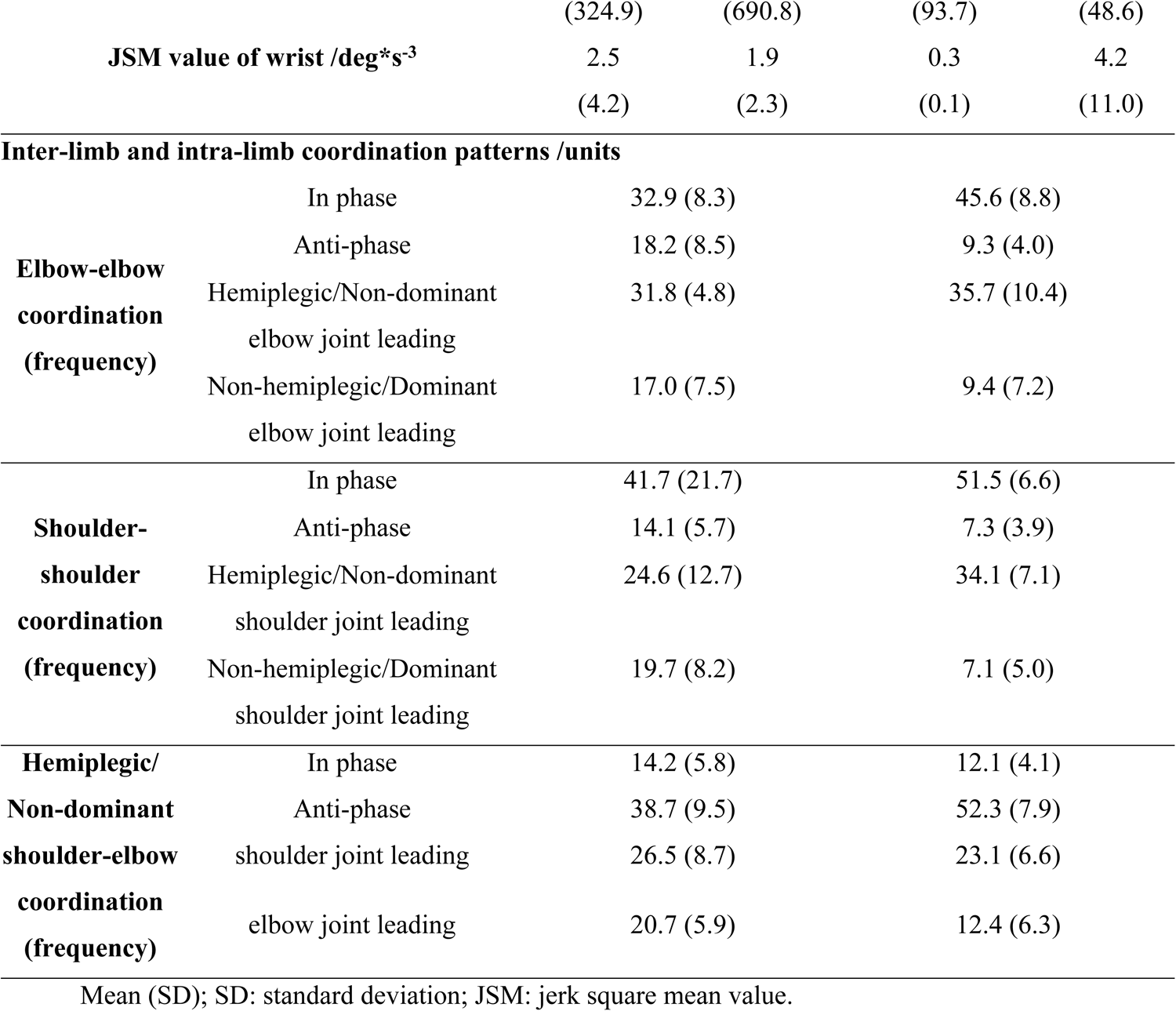
Kinematic variables between stroke and healthy group.

| Kinematic Variables | Stroke Group |  | Healthy Group |  |
| --- | --- | --- | --- | --- |
|  | (n = 17) |  | (n = 16) |  |
|  | Hemiplegic | Non-hemiplegic | Non-dominant (Left) | Dominant (Right) |
| Time and Peak velocity /units |  |  |  |  |
| Movement time /s | 45.3 (26.9) |  | 17.4 (5.9) |  |
| Movement initiation delay /s | 0.3 (0.5) |  | 0.1 (0.1) |  |
| Max elbow velocity /ms<sup>-1</sup> | 174.2<br>(110.8) | 128.0<br>(87.1) | 181.8<br>(40.9) | 164.4<br>(49.6) |
| Max shoulder velocity/ms<sup>-1</sup> | 127.1<br>(61.6) | 107.2<br>(63.2) | 213.3<br>(24.8) | 137.5<br>(55.0) |
| Compensatory Trunk Displacement /units |  |  |  |  |
| Total displacement of trunk/m | 0.6 (0.2) |  | 0.2 (0.1) |  |
| Max displacement of trunk/m | 0.1 (0.1) |  | 0.1 (0.1) |  |
| Smoothness – JSM value (JSM increased, smoothness decreased) /units |  |  |  |  |
| JSM value of elbow /deg*s<sup>-3</sup> | 1131.5<br>(940.1) | 810.5<br>(2502.9) | 1930.6<br>(913.3) | 884.7<br>(391.8) |
| JSM value of shoulder /deg*s<sup>-3</sup> | 322.3 | 237.0 | 116.5 | 67.1 |

|  |  |  |  |  |  |
| --- | --- | --- | --- | --- | --- |
|  |  | (324.9) | (690.8) | (93.7) | (48.6) |
|  | **JSM value of wrist /deg\*s<sup>-3</sup>** | 2.5 | 1.9 | 0.3 | 4.2 |
|  |  | (4.2) | (2.3) | (0.1) | (11.0) |
| **Inter-limb and intra-limb coordination patterns /units** |  |  |  |  |  |
| **Elbow-elbow**<br>**coordination**<br>**(frequency)** | In phase | 32.9 (8.3) |  | 45.6 (8.8) |  |
|  | Anti-phase | 18.2 (8.5) |  | 9.3 (4.0) |  |
|  | Hemiplegic/Non-dominant<br>elbow joint leading | 31.8 (4.8) |  | 35.7 (10.4) |  |
|  | Non-hemiplegic/Dominant<br>elbow joint leading | 17.0 (7.5) |  | 9.4 (7.2) |  |
| **Shoulder-**<br>**shoulder**<br>**coordination**<br>**(frequency)** | In phase | 41.7 (21.7) |  | 51.5 (6.6) |  |
|  | Anti-phase | 14.1 (5.7) |  | 7.3 (3.9) |  |
|  | Hemiplegic/Non-dominant<br>shoulder joint leading | 24.6 (12.7) |  | 34.1 (7.1) |  |
|  | Non-hemiplegic/Dominant<br>shoulder joint leading | 19.7 (8.2) |  | 7.1 (5.0) |  |
| **Hemiplegic/**<br>**Non-dominant**<br>**shoulder-elbow**<br>**coordination**<br>**(frequency)** | In phase | 14.2 (5.8) |  | 12.1 (4.1) |  |
|  | Anti-phase | 38.7 (9.5) |  | 52.3 (7.9) |  |
|  | shoulder joint leading | 26.5 (8.7) |  | 23.1 (6.6) |  |
|  | elbow joint leading | 20.7 (5.9) |  | 12.4 (6.3) |  |
| Mean (SD); SD: standard deviation; JSM: jerk square mean value. |  |  |  |  |  |

**Table 4.**
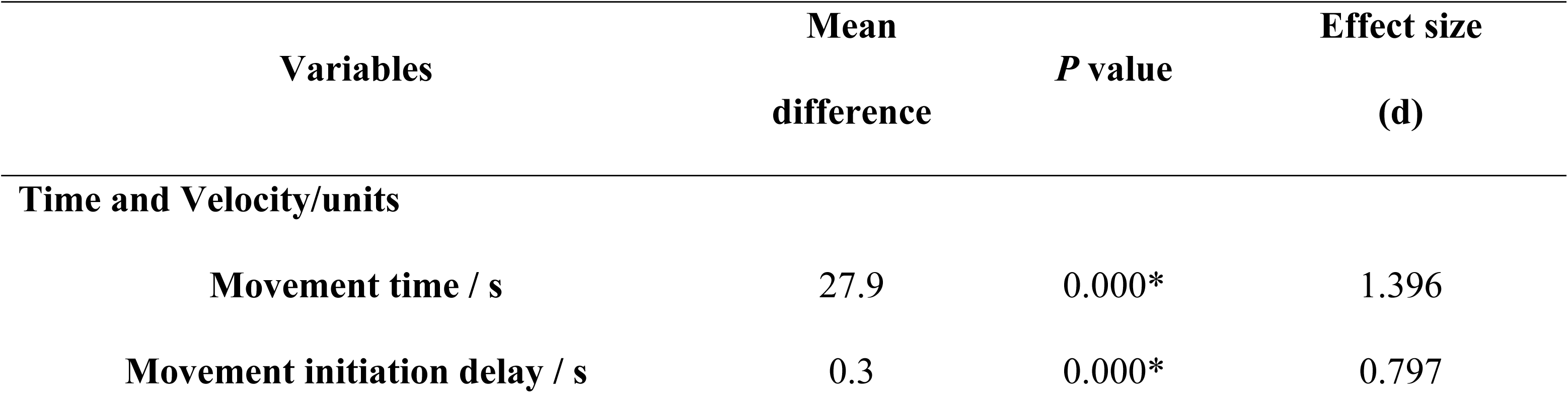
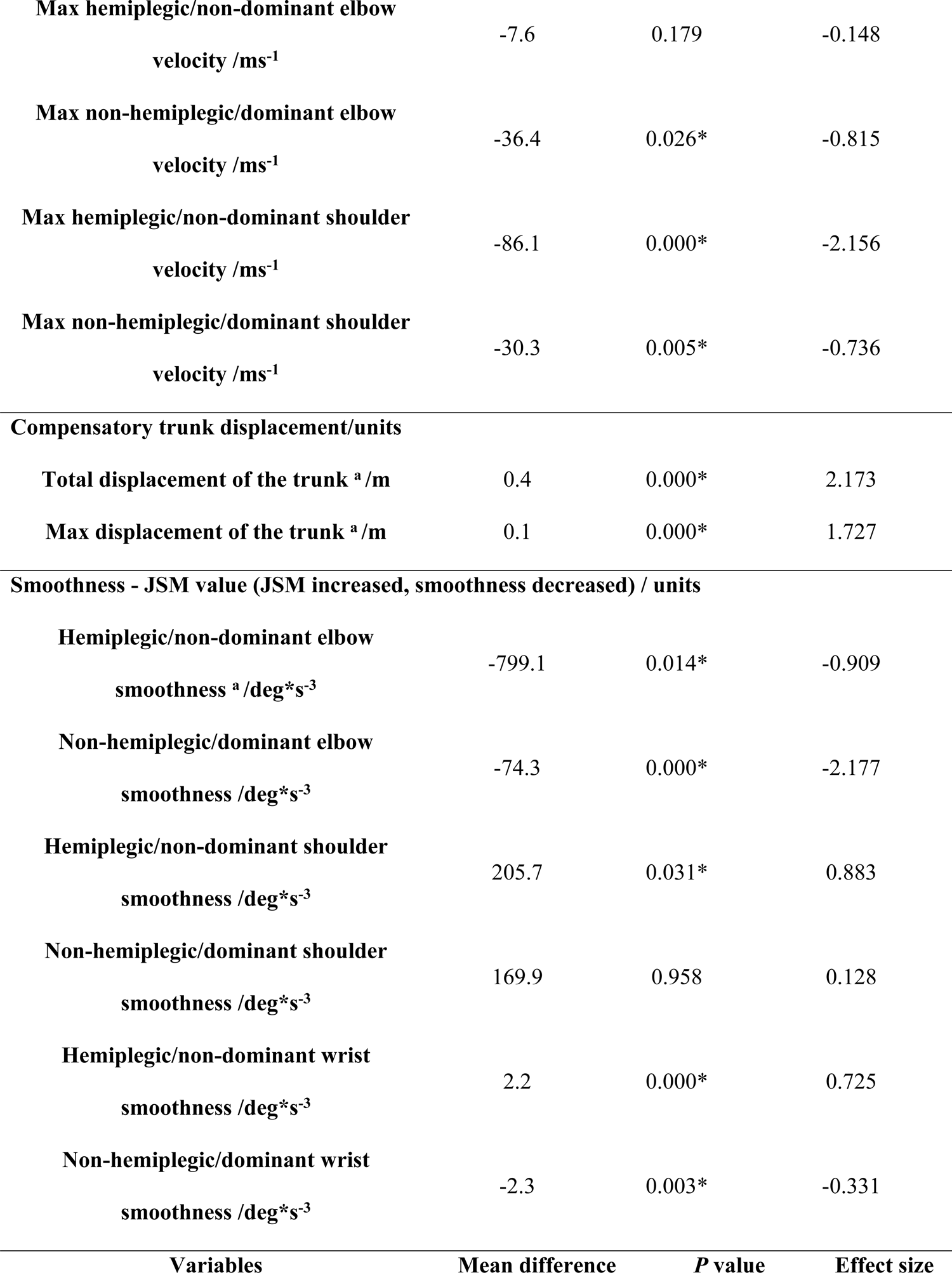
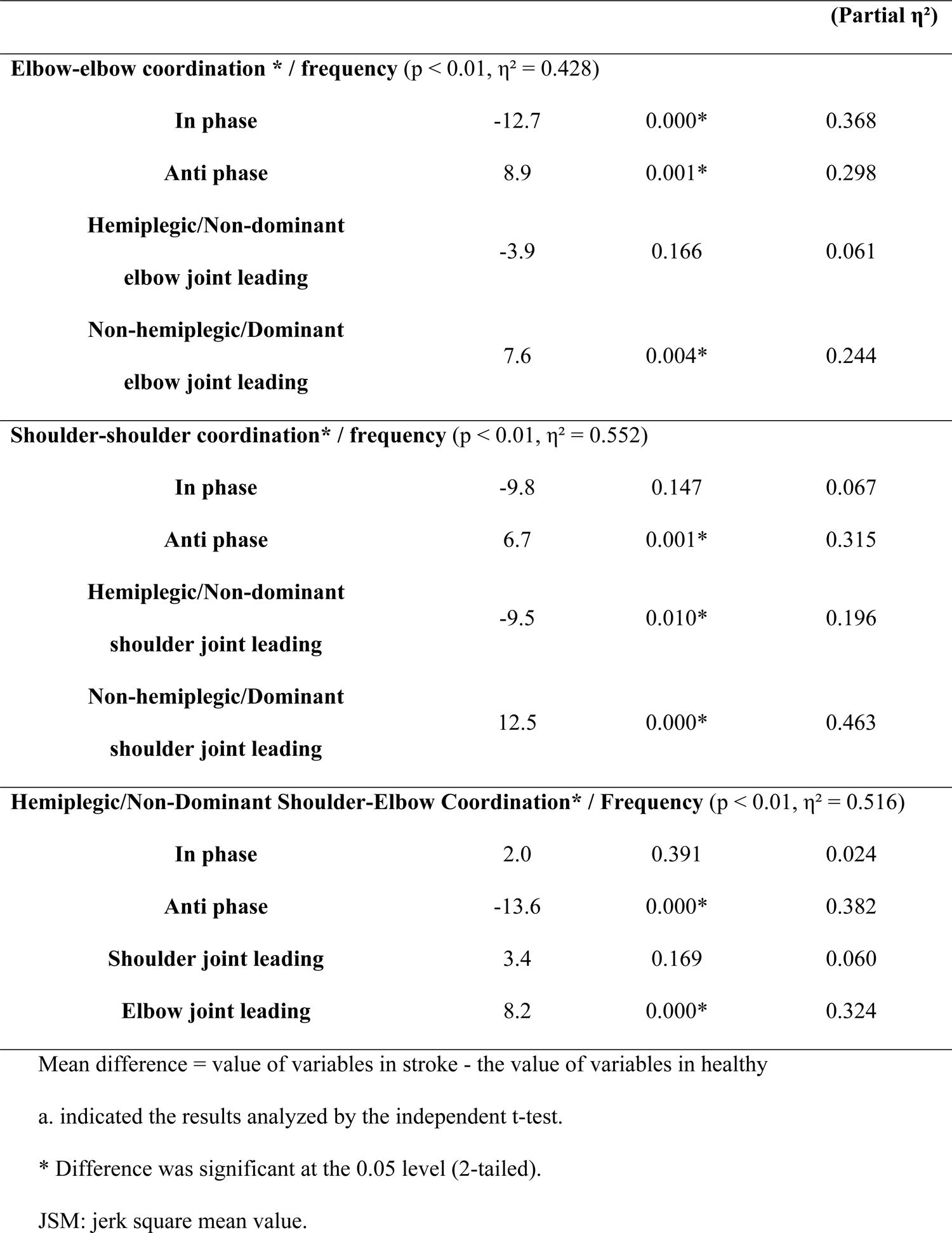
Difference of kinematic variables between stroke and healthy group during the folding towel task.

#### Compensatory Trunk Displacement

Compared to the healthy group, the stroke group exhibited notably larger total and peak trunk displacements (mean difference = 36.0 mm; *P* < .001; Cohen’s d = 2.173 and mean difference = 7.7 mm; *P* = .014; Cohen’s d = −0.909, respectively) (Tables 3 and 4).

#### Smoothness

We found that compared to the healthy group, the stroke group showed significantly increased JSM values (i.e., decreased smoothness) in the bilateral shoulder (mean difference = 205.7 deg*s^-3^; *P* = .031; Cohen’s d = .883 and mean difference = 169.9 deg*s^-3^; *P* = .958; Cohen’s d = .128, respectively), and hemiplegic wrist joints (mean difference = 2.2 deg*s^-3^; *P* < .001; Cohen’s d = .725). However, compared to the healthy group, we found that the stroke group had notably decreased JSM values in bilateral elbow (mean difference = −799.1 deg*s^-3^; *P* = .014; Cohen’s d = −.909 and mean difference = −74.3 deg*s^-3^; *P* < .001; Cohen’s d = −2.177, respectively) and non-hemiplegic wrist (mean difference = −2.3 deg*s^-3^; *P* = .003; Cohen’s d = −.331) (Tables 3 and 4).

#### Inter-limb and Intra-limb Coordination

We found significantly different coordination patterns between the two groups (Tables 3 and 4 and Fig 4). Regarding the bilateral elbow-elbow coordination pattern frequency (Fig 4a), we observed that compared to the healthy group, the stroke group showed significantly decreased frequency distribution for the in-phase (mean difference = −12.7; *P* < .001; partial η² = .368) and increased frequency distribution for the anti-phase and the non-hemiplegic elbow dominancy (mean difference = 8.9; *P* = .001; partial η² = .298 and mean difference = 7.6; *P* = .004; partial η² = .244, respectively). Regarding the bilateral shoulder-shoulder coordination pattern frequency (Fig 4b), we observed that compared to the healthy group, the stroke group showed significantly decreased frequency distribution for the hemiplegic shoulder leading (mean difference = −9.5; *P* = .010; partial η² = .196) and increased frequency distribution for the anti-phase and the non-hemiplegic shoulder dominancy (mean difference = 6.7; *P* = .001; partial η² = .315 and mean difference = 12.5; *P* < .001; partial η² = .463, respectively). For the hemiplegic shoulder-elbow coordination pattern frequency (Fig 4c), we observed that compared to the healthy group, the stroke group showed significantly decreased frequency distribution for the anti-phase (mean difference = −13.6; *P* < .001; partial η² = .382) and increased frequency distribution for the elbow dominancy (mean difference = 8.2; *P* < .001; partial η² = .324).

**Fig 4.**
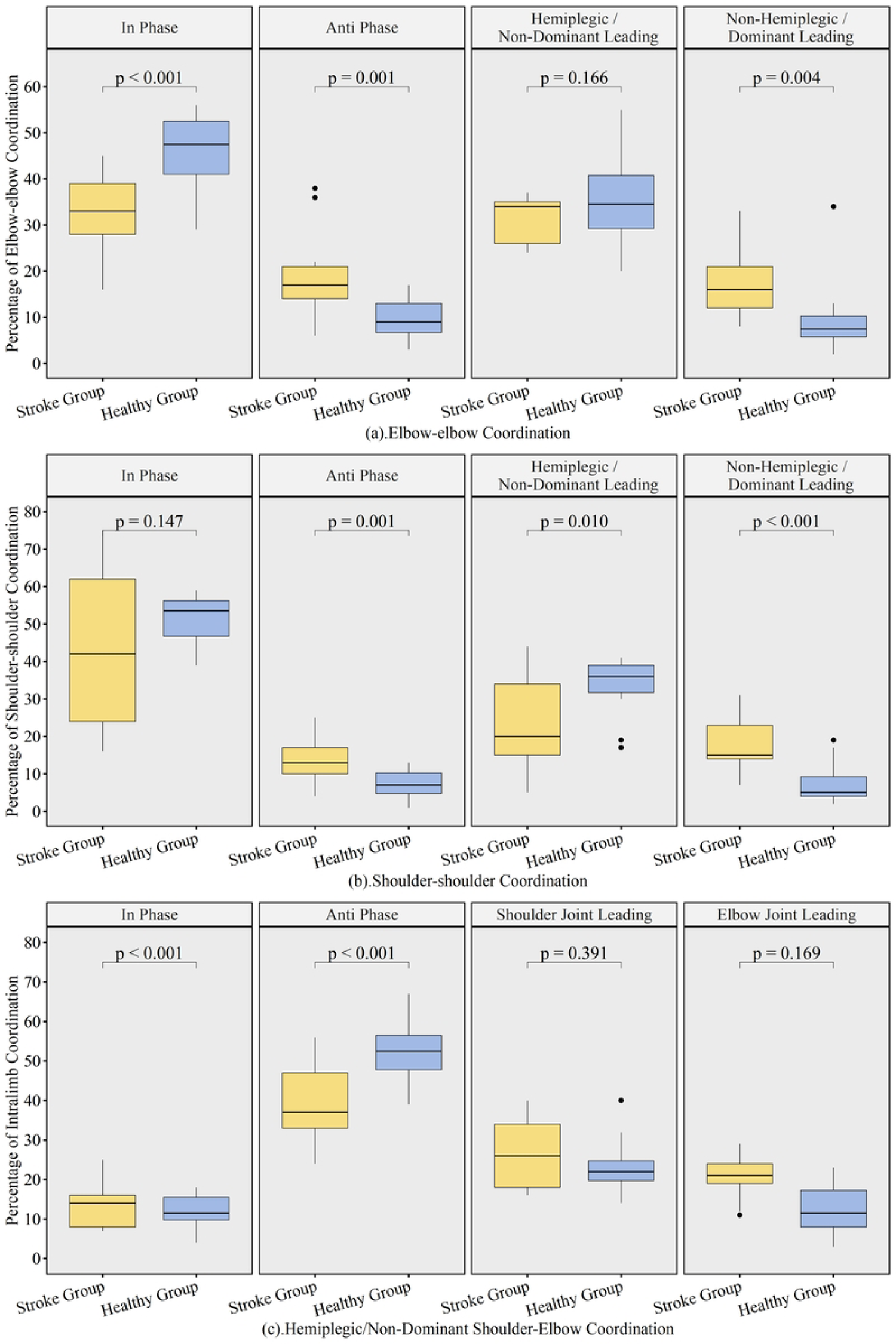
Differences in coordination pattern frequency between stroke and healthy group. (a). Elbow-elbow; (b) Shoulder-shoulder; (c) Hemiplegic shoulder-elbow.

#### Correlation coefficient

In the stroke group, we observed statistically significant positive associations between UE-FMA and movement-quality kinematic variables (Table 5). Specifically, the JSM value of the bilateral elbow decreased when UE-FMA score increased (r = −.500; *P* = .039 and r = −.670; *P* = .004, respectively). The JSM value of the hemiplegic shoulder and wrist decreased when UE-FMA score increased (r = −.540; *P* = .024 and r = −.520; *P* = .033, respectively). Additionally, stroke individuals showed greater hemiplegic shoulder-elbow coordination as UE-FMA score increased (r = .600; *P* = .010).

**Table 5.** Correlation coefficient between kinematic variables and clinical assessment in stroke group.

| **Kinematic Variables** | **Clinical Assessment** | **Correlation Coefficient [95%CI]** | **P value** |
| --- | --- | --- | --- |
| **Time and Velocity** |  |  |  |
| **Movement time** | UE-FMA | -0.320 [-0.694, 0.190] | 0.217 |
|  | ARAT | -0.350 [-0.711, 0.157] | 0.174 |
| **Movement initiation delay** | UE-FMA | -0.340 [-0.705, 0.168] | 0.179 |
|  | ARAT | -0.600 [-0.839, -0.168] | 0.010* |
| **Max hemiplegic elbow velocity** | UE-FMA | -0.130 [-0.575, 0.374] | 0.607 |
|  | ARAT | 0.210 [-0.301, 0.627] | 0.425 |
| **Max non-hemiplegic elbow velocity** | UE-FMA | -0.038 [-0.509, 0.451] | 0.884 |
|  | ARAT | 0.230 [-0.282, 0.640] | 0.370 |
| **Max hemiplegic shoulder velocity** | UE-FMA | -0.260 [-0.659, 0.252] | 0.311 |
|  | ARAT | -0.130 [-0.575, 0.374] | 0.607 |
| **Max non-hemiplegic shoulder velocity** | UE-FMA | -0.060 [-0.525, 0.433] | 0.818 |
|  | ARAT | 0.230 [-0.282, 0.640] | 0.384 |
| **Compensatory trunk displacement** |  |  |  |
| **Max displacement** | UE-FMA | 0.240 [-0.272, 0.646] | 0.355 |
|  | ARAT | -0.022 [-0.497, 0.464] | 0.933 |
| **Total displacement** | UE-FMA | 0.074 [-0.422, 0.536] | 0.778 |
|  | ARAT | 0.079 [-0.417, 0.539] | 0.763 |
| **Smoothness - JSM value (i.e., JSM increased, smoothness decreased)** |  |  |  |
| **Hemiplegic elbow smoothness** | UE-FMA | -0.500 [-0.791, -0.025] | 0.039* |
|  | ARAT | -0.190 [-0.615, 0.320] | 0.456 |
| **Non-hemiplegic elbow smoothness** | UE-FMA | -0.670 [-0.870, -0.280] | 0.004* |
|  | ARAT | -0.650 [-0.861, -0.246] | 0.005* |
| **Hemiplegic shoulder smoothness** | UE-FMA | -0.540 [-0.810, -0.080] | 0.024* |
|  | ARAT | -0.240 [-0.646, 0.272] | 0.357 |
| **Non-hemiplegic shoulder smoothness** | UE-FMA | -0.240 [-0.646, 0.272] | 0.345 |
|  | ARAT | -0.026 [-0.500, 0.460] | 0.921 |
| **Hemiplegic wrist smoothness** | UE-FMA | -0.520 [-0.800, -0.052] | 0.033* |
|  | ARAT | -0.360 [-0.717, 0.146] | 0.152 |
| **Non-hemiplegic wrist**<br>**smoothness** | UE-FMA | -0.370 [-0.722, 0.135] | 0.147 |
|  | ARAT | -0.290 [-0.676, 0.222] | 0.261 |
| **Inter-limb and intra-limb coordination** |  |  |  |
| **DOT: elbow-elbow** | UE-FMA | 0.380 [-0.123, 0.728] | 0.137 |
|  | ARAT | 0.430 [-0.064, 0.755] | 0.085 |
| **DOT: shoulder-shoulder <sup>a</sup>** | UE-FMA | -0.025 [-0.500, 0.461] | 0.925 |
|  | ARAT | 0.038 [-0.451, 0.509] | 0.884 |
| **DOT: hemiplegic/left shoulder-**<br>**elbow <sup>a</sup>** | UE-FMA | 0.600 [0.168, 0.840] | 0.010* |
|  | ARAT | 0.390 [-0.112, 0.733] | 0.126 |

UE-FMA: The Upper Extremity scores of the Fugl-Meyer Assessment; ARAT: the Action Research Arm Test. DOT: the unit vector scalar product.

Negative associations of ARAT with both the JSM values of the non-hemiplegic elbow (r = −.650; *P* = .005) and movement initiation delay (r = −.600; *P* = .010) were observed.

No significant relationships of clinical parameters with both trunk displacements and velocity were observed. Additionally, there was no notable association between UE-FMA and interlimb coordination patterns and between ARAT and inter-/intra-limb coordination patterns (Table 5).

#### Hand velocity undergoing folding the towel

In both healthy individuals using their non-dominant hand and stroke survivors using their hemiplegic hand, a similar pattern of variation in hand speed was observed while folding a towel (Fig 5). The hand speed changed throughout the task, increasing and then decreasing at each phase. The highest hand speed for both healthy individuals and stroke survivors was during Phase e of the task. Furthermore, the hemiplegic hand had a more frequent and limited range of speed changes during task completion compared to the non-dominant hand where the maximum speed change was 31.6% of that of healthy individuals (Figs 5a and 5b).

**Fig 5.**
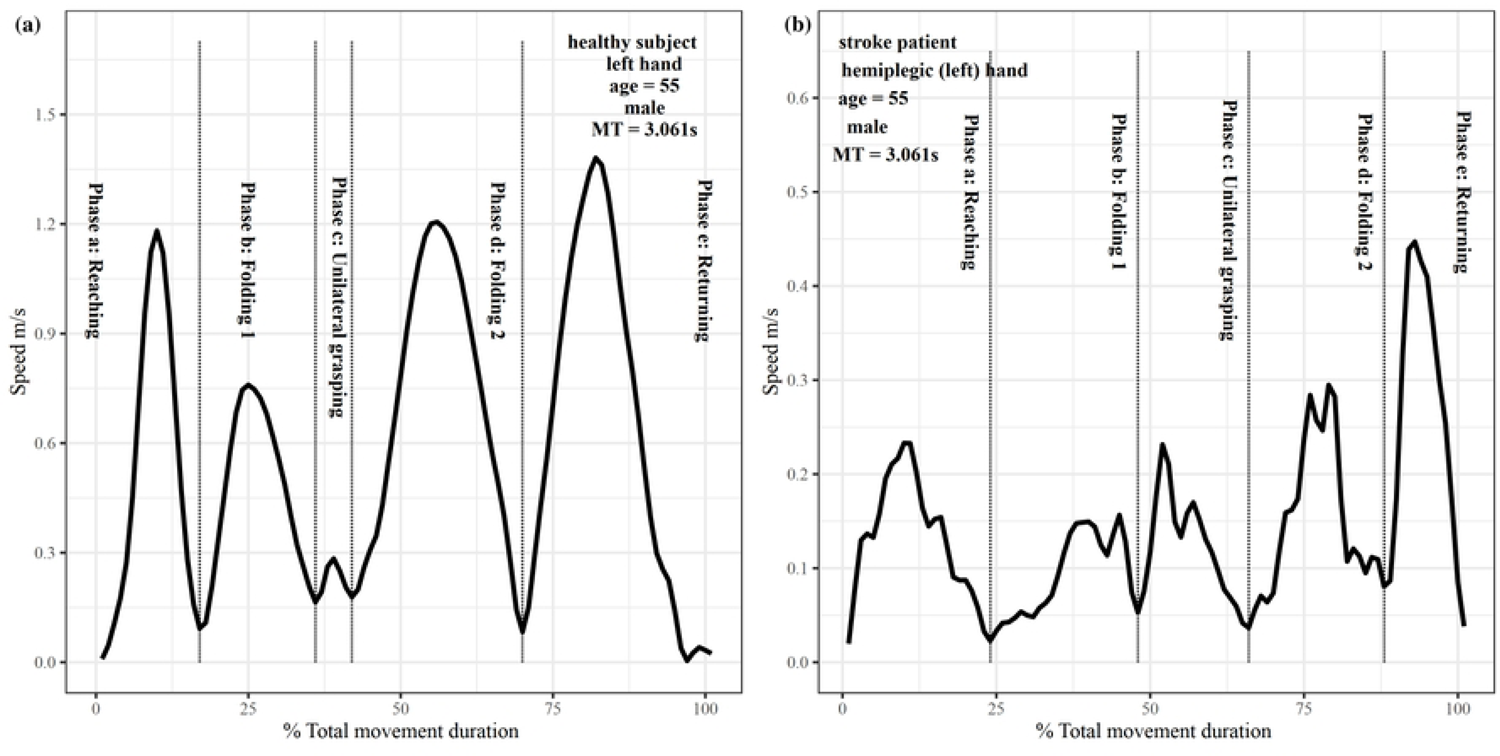
Curves of hand velocity variation throughout the total movement time in one stroke and healthy individual. Phase a: Reaching the towel with bilateral hands; Phase b: The first fold, folding the towel with bilateral hands from the anterior to posterior; Phase c: The unilateral hand grasping the same side corner of the towel (i.e., the hemiplegic hand for stroke patients; the non-dominant hand for healthy controls); Phase d: The second fold, folding the towel to the opposite side with the selected unilateral hand; Phase e: Returning the hand to the initial position. **(a)** Curves of non-dominant hand velocity variation throughout the total movement time in one representative healthy individual; **(b)** Curves of hemiplegic hand velocity variation throughout the total movement time in one representative stroke individual.

## Discussion

Our results showed that stroke individuals had longer time, slower initiation, lower max velocity, greater trunk displacement, fewer smoothness movements in the bilateral shoulder and hemiplegic wrist, and different coordination patterns compared to healthy controls. Additionally, we found that the UE-FMA was positively correlated with smoothness and intra-limb coordination, while the ARAT was negatively correlated with initiation delay in people with stroke.

### Difference in time and peak velocity between the two groups

Previous evidence has shown that movements in stroke individuals were slower compared to healthy individuals when performing unilateral tasks (17, 43). Our study extends these findings to bilateral tasks in stroke individuals, suggesting that movement time is effective for distinguishing between post-acute stroke and healthy individuals.

We found that stroke individuals exhibited similar elbow velocity on the hemiplegic side during bilateral tasks compared to healthy individuals. This finding was partly in line with previous evidence. Specifically, Hussain et al. (2018) documented that compared to control, stroke individuals with moderate impairment, not mild, displayed a significantly decreased peak velocity (43). It implies that the measure’s sensitivity varies with the severity of impairment.

### Compensatory trunk displacement

One notable distinction in movement patterns between the two groups was that stroke survivors tended to bend their trunks to fold a towel that was well within their arm’s reach (44). This compensatory strategy has been reported in earlier studies in unilateral reaching tasks (17, 19, 22). One possible explanation for this strategy is that the trunk may be controlled by the bilateral CST and therefore less impaired than the arm, which is predominantly controlled by the unilateral CST (45, 46). Another possible explanation is that the subject may fix the shoulder girdle to the trunk in an attempt to stabilize the unstable upper limb, reducing the number of degrees of freedom related to the upper limb. This could be supported by other findings of reduced variability in elbow and shoulder movements in stroke individuals with severe impairment (19). The additional trunk recruitment may supplant the reduced shoulder and elbow movement and provide the system additional stability. Another possibility was that the trunk muscles were recruited first to contribute to the task because of a lower threshold (47).

### Difference in smoothness between the two groups

Our findings suggest that differences in movement smoothness between stroke and healthy individuals vary by joints. Previous studies have shown worse smoothness in the hemiplegic upper limb during drinking tasks in stroke individuals compared to healthy individuals (17, 43, 48). Consistent with these findings, we observed decreased movement smoothness in the hemiplegic wrist and bilateral shoulders compared to healthy controls (49, 50), likely due to impaired motor control from muscle weakness and spasticity (51).

Interestingly, we found increased smoothness in the non-hemiplegic wrist and bilateral elbows compared to healthy controls. This might be because the longer movement time in stroke participants resulted in smaller JSM values for non-hemiplegic joints. Additionally, the simplicity of elbow movements in the task may have contributed to relatively intact smoothness in the hemiplegic elbow. However, the variability in task performance among participants means these findings should be interpreted cautiously.

### Differences in the frequency distribution in inter-limb and intra-limb coordination between the two groups

Previous studies used vector coding techniques to explore coordination patterns of bilateral lower limbs during walking in healthy individuals (37, 38). Our study is the first to apply this technique to bilateral upper limbs in stroke individuals. Our findings revealed that stroke individuals exhibited notable increases in anti-phase and non-hemiplegic dominancy in bilateral coordination patterns compared to healthy individuals. Previous studies and our study have observed movement initiation delay between the hemiplegic and non-hemiplegic side of stroke individuals during functional tasks (52, 53). Additionally, stroke individuals often compensate for their hemiplegic upper limb by using their non-hemiplegic limb (7). It implies that increased frequency for anti-phase and non-hemiplegic leading may aid in performing bilateral occupational activities in people with stroke.

Additionally, our results suggest that individuals with stroke tended to exhibit increases in hemiplegic elbow leading and decreases in anti-phase in intra-limb coordination patterns compared to healthy individuals. This aligns well with the previous study that found significantly decreased anti-phase in stroke individuals than healthy individuals (17). It is plausible that damage in CST made the ability to voluntarily move one segment independently of other segments dramatically decrease in stroke individuals (54–56). For instance, stroke individuals used the abnormal synergistic shoulder and elbow coupling to compensate for insufficient isolated shoulder flexion for adaptively completing reaching tasks, compared with healthy individuals (22). Further, Cirstea and Levin (2000) found that the elbow movement was initiated first during activities compared to the hemiplegic upper limb. This implies that enhancement in elbow leading and limitation in the anti-phase pattern in intra-limb coordination pattern was an integral part of the bilateral functional activities to promote the activities for stroke individuals (19). Furthermore, they partly explained the difference in intralimb coordination between stroke and healthy individuals through limited active ranges of motion and changes in motor thresholds for a group of muscles (19).

### Correlations between kinematic variables and clinical parameters in stroke group

Our findings revealed that UE-FMA was positively associated with smoothness and ARAT was negatively associated with delayed initiation. These findings were consistent with previous studies (57, 58). For instance, a study reported that the UE-FMA score increased when the smoothness of the hemiplegic hand increased during unilateral tasks (57). Further, Alt Murphy et al. (58) found that ARAT was notably correlated with movement time in stroke individuals during drinking tasks. Our results confirmed and extended these findings to bilateral functional tasks. These findings further support the notion that temporal kinematic variables were associated with clinical parameters involving the time component for task completion and movement quality-related kinematic variables were linked to clinical parameters involving the ability to perform joint movement (59).

Of note, our results suggest that UE-FMA was strongly related to intra-limb coordination patterns rather than bilateral coordination patterns in stroke individuals, likely due to UE-FMA mainly assesses hemiplegic upper limb motor function involving both multiple joints synergic and isolated movements (29). It implies that intra-limb may be a valuable tool for evaluating stroke movement quality.

No notable relationship between ARAT and inter/intra-limb coordination was observed, possibly because ARAT mainly focuses on task completion within a limited time and performance quality (33, 58). These findings suggest that inter/intra-limb coordination was indicative of changes in movement patterns and movement quality (i.e., impairment) rather than time and performance quality (i.e., activity).

## Limitations

There are several limitations to consider. Firstly, the small sample size of stroke and healthy individuals limits the generalizability of the results. Future studies should include a larger number of participants to enhance the applicability of the findings to the broader stroke population. Secondly, the accuracy of shoulder movement analysis is a concern. The shoulder is a complex joint, and there is debate over the best methods to calculate the centers of the glenohumeral, acromioclavicular, and sternoclavicular joints, as well as the definition of the 0-degree position and the computation of the range of motion (60). Thirdly, the study did not extract kinematic variables related to shoulder abduction-adduction during the towel-folding task. People with stroke often compensate with shoulder abduction in upper limb tasks, which limits the accuracy of the coordination pattern analysis between bilateral upper limbs and within the hemiplegic upper extremity.

## Conclusions

This study deepens the understanding of the folding towel task, a component of the Wolf Motor Function assessment, by identifying kinematic features linked to motor impairments in people with stroke. Kinematic assessments of interlimb and intralimb coordination prove sensitive in detecting differences in coordination patterns between people with stroke and healthy individuals during bilateral functional tasks. These metrics offer a precise quantification of the quality of bilateral coordination performance in people with stroke.

## Ethics approval and consent to participate

The study was conducted in accordance with the Declaration of Helsinki and approved by the Institutional Review Board of the Hong Kong Polytechnic University (HSEARS20220125002). Informed consent was obtained from all subjects involved in the study.

## Consent for publication

Written informed consent has been obtained from the patient(s) to publish this paper.

## Availability of data and materials

The data are held in a public repository (https://figshare.com/s/c6f94b5f957382f6d9a1)

## Authors’ contributions

Conceptualization: KWH, AS, and JYW.

Methodology: KWH and JYW.

Software: KWH and JYW.

Formal analysis: KWH and JYW.

Investigation: JYW, JWZ, and YNL.

Writing—original draft preparation: JYW, JJQZ, and JWZ.

Writing—review and editing: KWH, AS, FNK, and JJQZ.

Visualization: KWH and JYW.

Supervision: KWH.

Project administration: KWH.

Funding acquisition: KWH.

## Data Availability

data are held in a public repository (https://figshare.com/s/c6f94b5f957382f6d9a1)

## Acknowledgments

The authors thank all participants for their valuable contributions. The authors also thank Ms. XU and Mr. GU for assistance in the kinematic data processing.

## References

1. Gittler M, Davis AM. Guidelines for adult stroke rehabilitation and recovery. Jama. 2018;319(8):820–1.

2. Yang CL, Lim SB, Peters S, Eng JJ. Cortical Activation During Shoulder and Finger Movements in Healthy Adults: A Functional Near-Infrared Spectroscopy (fNIRS) Study. Front Hum Neurosci. 2020;14:260.

3. Liu F, Chen C, Bai Z, Hong W, Wang S, Tang C. Specific subsystems of the inferior parietal lobule are associated with hand dysfunction following stroke: A cross-sectional resting-state fMRI study. CNS Neurosci Ther. 2022.

4. Nguyen VT, Lu YH, Wu CW, Sung PS, Lin CC, Lin PY, et al. Evaluating interhemispheric synchronization and cortical activity in acute stroke patients using optical hemodynamic oscillations. J Neural Eng. 2022;19(3):36034.

5. Scano A, Guanziroli E, Mira RM, Brambilla C, Tosatti LM, Molteni F. Biomechanical assessment of the ipsilesional upper limb in post-stroke patients during multi-joint reaching tasks: A quantitative study. Frontiers in Rehabilitation Sciences. 2022;3:943397.

6. Culham JC, Cavina-Pratesi C, Singhal A. The role of parietal cortex in visuomotor control: what have we learned from neuroimaging? Neuropsychologia. 2006;44(13):2668–84.

7. Jones TA. Motor compensation and its effects on neural reorganization after stroke. Nature Reviews Neuroscience. 2017;18(5):267–80.

8. Kantak SS, Zahedi N, McGrath RL. Task-Dependent Bimanual Coordination After Stroke: Relationship With Sensorimotor Impairments. Arch Phys Med Rehabil. 2016;97(5):798–806.

9. Schaffer JE, Maenza C, Good DC, Przybyla A, Sainburg RL. Left hemisphere damage produces deficits in predictive control of bilateral coordination. Exp Brain Res. 2020;238(12):2733–44.

10. Rohrer B, Fasoli S, Krebs HI, Hughes R, Volpe B, Frontera WR, et al. Movement smoothness changes during stroke recovery. Journal of neuroscience. 2002;22(18):8297–304.

11. Caimmi M, Carda S, Giovanzana C, Maini ES, Sabatini AM, Smania N, et al. Using kinematic analysis to evaluate constraint-induced movement therapy in chronic stroke patients. Neurorehabilitation and neural repair. 2008;22(1):31–9.

12. Wolf SL, Lecraw DE, Barton LA, Jann BB. Forced use of hemiplegic upper extremities to reverse the effect of learned nonuse among chronic stroke and head-injured patients. Exp Neurol. 1989;104(2):125–32.

13. Wolf SL, Catlin PA, Ellis M, Archer AL, Morgan B, Piacentino A. Assessing Wolf motor function test as outcome measure for research in patients after stroke. Stroke. 2001;32(7):1635–9.

14. Lin J-H, Hsu M-J, Sheu C-F, Wu T-S, Lin R-T, Chen C-H, et al. Psychometric comparisons of 4 measures for assessing upper-extremity function in people with stroke. Physical therapy. 2009;89(8):840–50.

15. Schwarz A, Kanzler CM, Lambercy O, Luft AR, Veerbeek JM. Systematic review on kinematic assessments of upper limb movements after stroke. Stroke. 2019;50(3):718–27.

16. Gladstone DJ, Danells CJ, Black SE. The fugl-meyer assessment of motor recovery after stroke: a critical review of its measurement properties. Neurorehabil Neural Repair. 2002;16(3):232–40.

17. Alt Murphy M, Willén C, Sunnerhagen KS. Kinematic variables quantifying upper-extremity performance after stroke during reaching and drinking from a glass. Neurorehabil Neural Repair. 2011;25(1):71–80.

18. Alt Murphy M, Murphy S, Persson HC, Bergström UB, Sunnerhagen KS. Kinematic Analysis Using 3D Motion Capture of Drinking Task in People With and Without Upper-extremity Impairments. J Vis Exp. 2018(133).

19. Cirstea MC, Levin MF. Compensatory strategies for reaching in stroke. Brain. 2000;123(5):940–53.

20. Carpinella I, Jonsdottir J, Ferrarin M. Multi-finger coordination in healthy subjects and stroke patients: a mathematical modelling approach. Journal of neuroengineering and rehabilitation. 2011;8(1):1–20.

21. Clark RA, Pua YH, Bryant AL, Hunt MA. Validity of the Microsoft Kinect for providing lateral trunk lean feedback during gait retraining. Gait Posture. 2013;38(4):1064–6.

22. Levin MF, Liebermann DG, Parmet Y, Berman S. Compensatory Versus Noncompensatory Shoulder Movements Used for Reaching in Stroke. Neurorehabil Neural Repair. 2016;30(7):635–46.

23. Liddy JJ, Zelaznik HN, Huber JE, Rietdyk S, Claxton LJ, Samuel A, et al. The efficacy of the Microsoft Kinect TM to assess human bimanual coordination. Behavior research methods. 2017;49(3):1030–47.

24. Pfister A, West AM, Bronner S, Noah JA. Comparative abilities of Microsoft Kinect and Vicon 3D motion capture for gait analysis. J Med Eng Technol. 2014;38(5):274–80.

25. Sakata K, Kogure A, Hosoda M, Isozaki K, Masuda T, Morita S. Evaluation of the age-related changes in movement smoothness in the lower extremity joints during lifting. Gait & posture. 2010;31(1):27–31.

26. Xu X, McGorry RW, Chou LS, Lin JH, Chang CC. Accuracy of the Microsoft Kinect for measuring gait parameters during treadmill walking. Gait Posture. 2015;42(2):145–51.

27. Kwakkel G, van Wegen EEH, Burridge JH, Winstein CJ, van Dokkum LEH, Alt Murphy M, et al. Standardized Measurement of Quality of Upper Limb Movement After Stroke: Consensus-Based Core Recommendations From the Second Stroke Recovery and Rehabilitation Roundtable. Neurorehabil Neural Repair. 2019;33(11):951–8.

28. Bernhardt J, Hayward KS, Kwakkel G, Ward NS, Wolf SL, Borschmann K, et al. Agreed definitions and a shared vision for new standards in stroke recovery research: the stroke recovery and rehabilitation roundtable taskforce. International Journal of Stroke. 2017;12(5):444–50.

29. Fugl Meyer AR, Jaasko L, Leyman I. The post stroke hemiplegic patient. I. A method for evaluation of physical performance. Scandinavian journal of rehabilitation medicine. 1975;7(1):13–31.

30. Kim H, Her J, Ko J, Park D-s, Woo J-H, You Y, et al. Reliability, concurrent validity, and responsiveness of the Fugl-Meyer Assessment (FMA) for hemiplegic patients. Journal of Physical Therapy Science. 2012;24(9):893–9.

31. Chen H-f, Lin K-c, Wu C-y, Chen C-l. Rasch validation and predictive validity of the action research arm test in patients receiving stroke rehabilitation. Archives of physical medicine and rehabilitation. 2012;93(6):1039–45.

32. Lang CE, Wagner JM, Dromerick AW, Edwards DF. Measurement of upper-extremity function early after stroke: properties of the action research arm test. Archives of physical medicine and rehabilitation. 2006;87(12):1605–10.

33. Lyle RC. A performance test for assessment of upper limb function in physical rehabilitation treatment and research. International journal of rehabilitation research. 1981;4(4):483–92.

34. Coats RO, Fath AJ, Astill SL, Wann JP. Eye and hand movement strategies in older adults during a complex reaching task. Exp Brain Res. 2016;234(2):533–47.

35. Nan HC, Rambely AS. Peak velocity of elbow joint during touching contra lateral shoulder activity for normal subject. AIP Conference Proceedings; April: AIP Publishing LLC; 2017. p. 020034.

36. Jafri M, Brown S, Arnold G, Abboud R, Wang W. Kinematical analysis of the trunk, upper limbs and fingers during minimal access surgery when using an armrest. Ergonomics. 2015;58(11):1868–77.

37. Needham R, Naemi R, Chockalingam N. Quantifying lumbar–pelvis coordination during gait using a modified vector coding technique. Journal of biomechanics. 2014;47(5):1020–6.

38. Needham RA, Naemi R, Chockalingam N. A new coordination pattern classification to assess gait kinematics when utilising a modified vector coding technique. Journal of biomechanics. 2015;48(12):3506–11.

39. Chang R, Van Emmerik R, Hamill J. Quantifying rearfoot–forefoot coordination in human walking. Journal of biomechanics. 2008;41(14):3101–5.

40. Sparrow W, Donovan E, Van Emmerik R, Barry E. Using relative motion plots to measure changes in intra-limb and inter-limb coordination. Journal of motor behavior. 1987;19(1):115–29.

41. Yap B, Sim C. Comparison of various types of normality tests. Journal of Statistical Computation and Simulation. 2011;81(12):2141–55.

42. Cohen J. Statistical power analysis for the behavioral sciences. 2nd ed. Hillsdale, N.J: L. Erlbaum Associates; 1988.

43. Hussain N, Murphy MA, Sunnerhagen KS. Upper limb kinematics in stroke and healthy controls using target-to-target task in virtual reality. Frontiers in neurology. 2018;9(MAY):300-.

44. Alt Murphy M, Häger CK. Kinematic analysis of the upper extremity after stroke–how far have we reached and what have we grasped? Physical Therapy Reviews. 2015;20(3):137–55.

45. Fujiwara T, Sonoda S, Okajima Y, Chino N. The relationships between trunk function and the findings of transcranial magnetic stimulation among patients with stroke. J Rehabil Med. 2001;33(6):249–55.

46. Karthikbabu S, Chakrapani M, Ganeshan S, Rakshith KC, Nafeez S, Prem V. A review on assessment and treatment of the trunk in stroke: A need or luxury. Neural Regen Res. 2012;7(25):1974–7.

47. Shaikh T, Goussev V, Feldman AG, Levin MF. Arm-trunk coordination for beyond-the-reach movements in adults with stroke. Neurorehabil Neural Repair. 2014;28(4):355–66.

48. Bustrén E-L, Sunnerhagen KS, Alt Murphy M. Movement kinematics of the ipsilesional upper extremity in persons with moderate or mild stroke. Neurorehabilitation and neural repair. 2017;31(4):376–86.

49. Lang CE, Wagner JM, Bastian AJ, Hu Q, Edwards DF, Sahrmann SA, et al. Deficits in grasp versus reach during acute hemiparesis. Exp Brain Res. 2005;166(1):126–36.

50. Nowak DA. The impact of stroke on the performance of grasping: usefulness of kinetic and kinematic motion analysis. Neurosci Biobehav Rev. 2008;32(8):1439–50.

51. Balkaya M, Cho S. Optimizing functional outcome endpoints for stroke recovery studies. J Cereb Blood Flow Metab. 2019;39(12):2323–42.

52. Jones RD, Donaldson IM, Parkin PJ. Impairment and recovery of ipsilateral sensory-motor function following unilateral cerebral infarction. Brain. 1989;112(1):113–32.

53. Chae J, Yang G, Park BK, Labatia I. Delay in initiation and termination of muscle contraction, motor impairment, and physical disability in upper limb hemiparesis. Muscle Nerve. 2002;25(4):568–75.

54. Dewald JP, Pope PS, Given JD, Buchanan TS, Rymer WZ. Abnormal muscle coactivation patterns during isometric torque generation at the elbow and shoulder in hemiparetic subjects. Brain. 1995;118(2):495–510.

55. Lang CE, Schieber MH. Reduced muscle selectivity during individuated finger movements in humans after damage to the motor cortex or corticospinal tract. J Neurophysiol. 2004;91(4):1722–33.

56. Lang CE, Bland MD, Bailey RR, Schaefer SY, Birkenmeier RL. Assessment of upper extremity impairment, function, and activity after stroke: foundations for clinical decision making. J Hand Ther. 2013;26(2):104–15.

57. van Dokkum L, Hauret I, Mottet D, Froger J, Métrot J, Laffont I. The contribution of kinematics in the assessment of upper limb motor recovery early after stroke. Neurorehabilitation and neural repair. 2014;28(1):4–12.

58. Alt Murphy M, Willén C, Sunnerhagen KS. Movement kinematics during a drinking task are associated with the activity capacity level after stroke. Neurorehabilitation and neural repair. 2012;26(9):1106–15.

59. Alt Murphy M, Resteghini C, Feys P, Lamers I. An overview of systematic reviews on upper extremity outcome measures after stroke. BMC neurology. 2015;15:1–15.

60. Blache Y, Martinez R, Dumas R, Begon M, Hagemeister N, Duprey S. Chapter 20 - Motion analysis and modeling of the shoulder: challenges and potential applications. In: Scataglini S, Paul G, editors. DHM and Posturography: Academic Press; 2019. p. 261–71.

